# A Neuroimaging-based Precision Medicine Framework for Depression

**DOI:** 10.1101/2022.10.21.22281154

**Authors:** Yao Xiao, Shuai Dong, Rongxin Zhu, Fay Y. Womer, Ran Zhang, Jingyu Yang, Luheng Zhang, Juan Liu M.D., Weixiong Zhang, Zhongchun Liu, Xizhe Zhang, Fei Wang

## Abstract

**Objective:** Developing a neuroimaging-based precision medicine framework for depression.

**Methods:** The study was conducted in two stages at two sites: development of a neuroimaging-based subtyping and precise repetitive transcranial magnetic stimulation (rTMS) strategy for depression at Center 1 and its clinical application at Center 2. Center 1 identified depression subtypes and subtype-specific rTMS targets based on amplitude of low frequency fluctuation (ALFF) in a cohort of 238 major depressive disorder patients and 66 healthy controls (HC). Subtypes were identified using a Gaussian Mixture Model, and subtype-specific rTMS targets were selected based on dominant brain regions prominently differentiating depression subtypes from HC. Subsequently, one classifier trained per Center 1 findings for subtyping and subtype-specific rTMS targets were employed to deliver two-week precise rTMS to 72 hospitalized, depressed youths at Center 2. MRI and clinical assessments were obtained at baseline, midpoint, and treatment completion for evaluation.

**Results:** Two neuroimaging subtypes of depression, archetypal and atypical depression, were identified based on distinct frontal-posterior functional imbalance patterns as measured by ALFF. The dorsomedial prefrontal cortex was identified as the rTMS target for archetypal depression, and the occipital cortex for atypical depression. Following precise rTMS, ALFF alterations were normalized in both archetypal and atypical depressed youths, corresponding with symptom response of 90.00% in archetypal depression and 70.73% in atypical depression.

**Conclusions:** A precision medicine framework for depression was developed based on frontal-posterior functional imbalance and implemented with promising results. Future randomized controlled trials are warranted.

Chinese Clinical Trial Registry identifier: ChiCTR2100045391

## INTRODUCTION

Depression is one of the most common psychiatric disorders with a lifetime prevalence of 15%-18% [1], and around 350 million people suffer from depression worldwide [2]. Depression severely impairs many aspects of daily functioning, including education and employment [3], economy [4], and intimate relationships [5], leading to stunted individual and social development. Unfortunately, the efficacy of conventional treatments for depression remains suboptimal; up to 40% of patients who receive antidepressant monotherapy achieve remission, even with a treatment duration of 3-6 months [6]. Moreover, in sharp contrast to many other medical conditions, the global burden of depression has not decreased in the past thirty years [7]. Altogether, these issues underscore the urgent need for novel and precise therapeutic strategies for depression.

Symptom-based diagnostic criteria result in notorious heterogeneity in depression, which impeded precision medicine [8]. One approach to eliminating the heterogeneity is to define depression into different subtypes [9]. Clinical specifiers are empirically applied by the Diagnostic and Statistical Manual of Mental Disorders, 5^th^ Edition (DSM-5) for subtype characterization [10], yet the descriptive symptom features fail to differentiate depression subtypes etiologically.

Neuroimaging studies have leveraged a tectonic shift in reconceptualizing depression beyond symptoms [8, 11, 12]. Combined with machine learning techniques, depression could be delineated into distinct neuroimaging subtypes with subtype-specific neural deficits in a data-driven fashion [13, 14]. The resulting reproducible and objective neurobiomarkers for subtyping would have highly promising potential to guide precision medicine in depression. In our prior work, we identified two neurobiological subtypes in psychiatric disorders including major depressive disorder (MDD) based on the amplitude of low-frequency fluctuations (ALFF): archetypal and atypical subtypes [15]. ALFF may reflect regional spontaneous neural activities at rest and has shown to be a reliable measure for psychiatric disorders [16, 17]. The identified archetypal and atypical subtypes had converse patterns of frontal-posterior functional imbalance associated with differentiation in white matter integrity, cortical thickness, polygenic risk scores, tissue profiles for risk gene expression, and medication effects, implicating frontal-posterior functional imbalance as a potential neuromodulation target for precise repetitive transcranial magnetic stimulation (rTMS) in depression [15].

As a non-invasive form of brain stimulation, rTMS has been approved by the U.S. Food and Drug Administration for adult treatment-resistant depression (TRD) in 2008 [18]. Studies adopting rTMS as first-line treatment in depressed youths have been rapidly increasing ever since, confirming good tolerability, safety, and efficacy in adolescent populations as well [19]. Compared with pharmacologic methods or electroconvulsive therapy, rTMS can directly target dysfunctional brain regions, and stimulation parameters can be adjusted accordingly [20]. Therefore, it is one of the most promising antidepressant therapies for achieving precision [21, 22]. However, the majority of studies in rTMS precision have aimed at improving spatial targeting of the dorsolateral prefrontal cortex (dlPFC), which is an empirically selected target and may not induce a response in all depressed individuals [23]. Prior study has shown that rTMS response varies significantly among neurophysiological depression subtypes [24]. Therefore, identification of subtype-specific rTMS targets based on neuroimaging subtypes of depression hold a great expectation in optimizing the precision of current rTMS therapy.

We aim at developing and implementing a neuroimaging-based subtyping and precise rTMS strategy for depression. Concerningly, depression during adolescence and early adulthood often results in significant developmental disruptions and has long-term implications for future psychiatric illness and functioning in adulthood [25]. Nevertheless, 30-50% of depressed adolescents do not respond to their first treatment intervention, and 10% of depressed adolescents do not improve despite multiple treatment trials [26]. Young populations with depression would benefit significantly from more effective and precise treatment. Hence, we implemented the novel treatment strategy in depressed youths. The study was conducted in two stages at two sites. We first recruited a cohort of MDD individuals and healthy controls (HC) at Center 1 to identify depression subtypes and subtype-specific rTMS targets based on ALFF patterns. We then employed a classifier trained per Center 1 findings for depression subtyping and identification of subtype-specific rTMS targets to deliver precise rTMS to a group of hospitalized, depressed youths at Center 2. Precise rTMS was performed with pre- and post-treatment assessments for functional MRI changes and symptomatic response.

## METHODS

The study was carried out at two sites, Renmin Hospital of Wuhan University (Center 1) and Affiliated Nanjing Brain Hospital of Nanjing Medical University (Center 2). The development of the neuroimaging-based subtyping and precise rTMS strategy for depression was conducted at Center 1. Precise rTMS and neuroimaging and symptomatic evaluations in hospitalized, depressed youths were performed at Center 2. (Figure 1)

**Figure 1.**
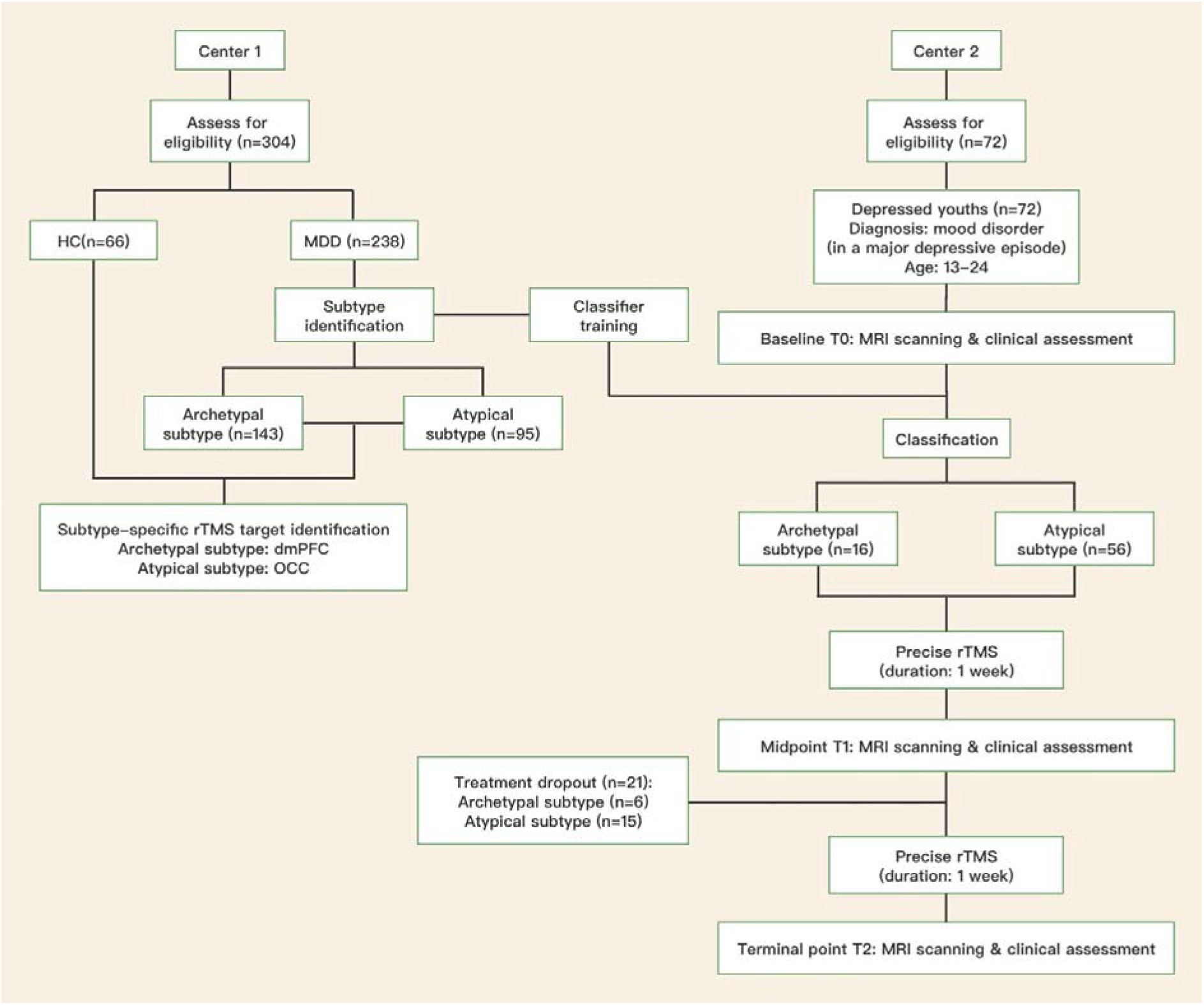
Schema of developing a neuroimaging-based subtyping and precise repetitive transcranial magnetic stimulation (rTMS) strategy for depression and implementing it in hospitalized, depressed youths. HC=healthy control; MDD=major depressive disorder; dmPFC=dorsomedial prefrontal cortex; OCC=occipital cortex.

### Participants

Center 1 participants consisted of 238 outpatients with DSM-IV diagnosis of MDD and 66 HC aged 13 to 55. HC did not have personal or family history of psychiatric disorders. Center 2 participants consisted of 72 hospitalized youths aged 13 to 24 with DSM-IV diagnosis of mood disorder (38 MDD and 34 bipolar II disorder) and were in a current major depressive episode and had a baseline score of at least 17 on the 17-item Hamilton Rating Scale for Depression (HAMD-17) or at least 16 on the 14-items Hamilton Rating Scale for Anxiety (HAMA). Participants were excluded at both centers if any major medical condition, neurological disorder, MRI contraindications, or excessive head motion during MRI scan. See Supplementary Material for additional participant characteristics.

All participants provided written informed consents; for those < 18 years, written informed consents were obtained from their legal guardians and assent from the minor participant. The precise rTMS trial was approved by the Ethics Committee of Renmin Hospital of Wuhan University and Nanjing Brain Hospital and registered in the Chinese Clinical Trial Registry (ChiCTR2100045391).

### MRI data acquisition and processing

Participants underwent 3.0 T MRI scanning at their respective centers. Preprocessing of resting-state functional MRI for ALFF analyses was the same across both centers. See Supplementary Material for center-specific protocols and data processing.

### Identifying neuroimaging subtypes of depression

For each MDD patient, the high-dimensional ALFF data were reduced to two dimensions by using the t-distributed Stochastic Neighbor Embedding algorithm [27]. The MDD patients were then divided into two subtypes based on the Gaussian Mixture Model algorithm [28]. The stability of clustering results was tested using the Consensus Clustering method [29]. The subtype diagnostic performances were further evaluated by using Support Vector Machine (SVM) classifiers to distinguish each subtype and total MDD sample from HC. Five-fold cross-validation was used to evaluate the performance of each SVM classifier; the F1 score and area under the curve (AUC) were calculated as measures. Permutation testing [30] was employed to further assess the reliability of subtypes with a significance level of p < 0.002. For detailed methods, see online Supplementary Material.

### Identifying subtype-specific rTMS targets

We separately extracted 90 brain regions’ ALFF values based on automated anatomical labeling-90 atlas [31]; for every brain region, an SVM classifier was trained to distinguish depression subtype from HC. Five-fold cross-validation was used to evaluate the classifier performance measured by F1 scores. For each subtype, we ranked the 90 brain regions based on their F1 scores and selected top-10 brain regions as candidate regions. Subsequently, the anatomical distribution of the top-10 brain regions was examined, and the dominant regions of alteration were identified. The current neurobiological understanding was then used to finalize an rTMS target for each subtype. See Supplementary Material for further details.

### Precise rTMS for depressed youths

Hospitalized, depressed youths at Center 2 received precise rTMS with a total of 20 sessions over two weeks. An SVM classifier was trained based on neuroimaging data of Center 1 for depression subtype separation. The classification model was then used to divide depressed youths into subtypes at Center 2. For the subtype labeled as ‘archetypal’, the stimulation was high frequency (10Hz), with 1,200 pulses per session; for the subtype labeled as ‘atypical’, the stimulation was low frequency (1Hz), with 1,000 pulses per session (see results section for label characteristics). Stimulation of both subtypes was applied at 100% resting motor threshold. The international 10-10 system and individualized three-dimensional MRI were jointly used for the precise localization of rTMS targets in each participant. For further details, see Supplementary Material.

### Neuroimaging data assessments

At Center 1, whole-brain voxel-wise two-sample t-test of ALFF values was performed between each subtype and HC to examine subtype ALFF alterations. At Center 2, whole-brain voxel-wise paired t-test was performed to examine ALFF alterations post- and pre-rTMS for each subtype. Voxel-level statistical significance was set to p < 0.05, with Gaussian random field multiple-comparison correction (cluster-level p < 0.05, cluster size > 250).

### Clinical assessments

Clinical assessments were performed by three blinded, certified psychiatrists at baseline T0 (one day before precise rTMS), midpoint T1 (after one-week precise rTMS), and terminal point T2 (after two-week precise rTMS), on the same days as participant MRI scans. The primary outcome was symptom severity as measured by both the HAMD-17 and HAMA. A secondary outcome was suicidality as measured by Item 3 on HAMD-17. The response was defined as ≥ 50% symptom reduction on scales and suicidality item; specifically, the primary outcome was considered overall response when HAMD-17 or HAMA reached response. Remission for primary and secondary outcomes was determined by HAMD-17 or HAMA scores ≤ 7, and a score of 0 for Item-3 of HAMD-17, respectively.

## RESULTS

### Identified depression neuroimaging subtypes reveal a distinct frontal-posterior functional imbalance

Using machine-learning techniques, we identified two neuroimaging subtypes in the Center 1 MDD cohort (n=238). The two depression subtypes (n_1_=143 and n_2_=95, respectively) showed distinct frontal-posterior functional imbalance patterns, specifically across the prefrontal and occipital cortices (Figure 2A). The ALFF patterns of the two subtypes were consistent with our previous finding [15], so we denoted the two subtypes identified herein as *archetypal depression* and *atypical depression*. Compared to HC, archetypal depression had significantly increased ALFF in frontal regions (prefrontal cortex, limbic, para-limbic, and striatum) and significantly decreased ALFF in posterior regions (primary visual, sensory, motor cortices and unimodal association cortices); whereas atypical depression exhibited significantly decreased ALFF in similar frontal regions and significantly increased ALFF in similar posterior regions. A clustering stability test showed that both subtypes had robust stability (m_1_=0.93, m_2_=0.73, Figure 2B). No significant difference was detected between two subtypes in terms of age (t=-0.68, p=0.50) or sex (χ^2^=0.11, p=0.74).

**Figure 2.**
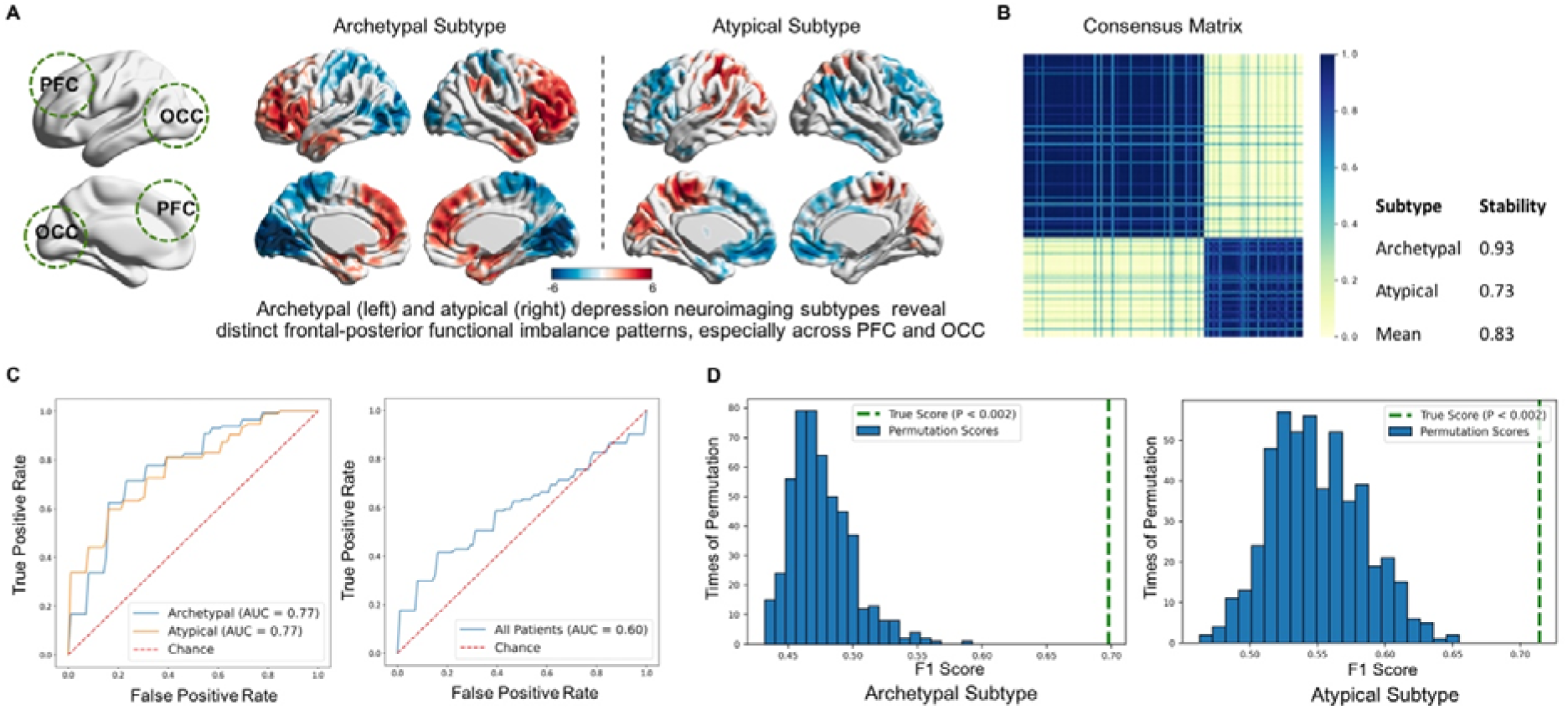
Identified depression neuroimaging subtypes reveal a distinct frontal-posterior functional imbalance. Panel A shows the neuroimaging characteristics of depression archetypal subtype and atypical subtype (whole-brain voxel-wise two-sample t-test of the amplitude of low frequency fluctuations between each subtype and HC with voxel-level statistical significance p < 0.05, followed by Gaussian random field multiple-comparison correction; cluster-level p < 0.05, cluster size > 250). Red represents t > 0, blue represents t < 0. Panel B shows the heat map of consensus matrix for testing clustering stability based on Consensus Clustering method. For each pair of patients in the matrix, the proportion of them being clustered to the same subtype in all 500 runs was presented through a color gradient set to 0-1; the dark color indicated the proportion to be high, and the light color the opposite. Both subtypes had good stability. Panel C shows the plotted ROC curves of classifiers trained for distinguishing each subtype and the total MDD sample from HC; both subtypes achieved higher AUC than total sample. Panel D shows results of permutation testing for assessing subtype reliability. For patients in each subtype, the subtype labels were randomly permuted into either archetypal or atypical. For each subtype, one classifier was trained for dividing the randomly labeled ‘subtype’ from HC and F1 score was calculated; the process repeated 500 times. True F1 scores exceeded 95% confidence interval of permutated F1 scores in both subtypes, suggesting permutation testing to be statistically significant at p < 0.002. Both subtypes achieved great reliability. PFC=prefrontal cortex; OCC=occipital cortex; ROC= receiver operating characteristic; MDD=major depressive disorder; HC=healthy control; AUC=area under the curve.

Remarkably, both subtypes had superior diagnostic capability compared to that of the total MDD sample (Figure 2C). We trained three SVM classifiers based on ALFF to separately distinguish the total MDD group and the archetypal and atypical subtypes from HC. Classifiers for the archetypal and atypical depression collectively achieved higher performance (archetypal: F1=0.70, AUC=0.77; atypical: F1=0.71, AUC=0.77) than that of MDD diagnosis (F1=0.44, AUC=0.60), supporting the validity of the two identified depression subtypes. The statistical significance of permutation testing confirmed the reliability of the two subtypes (p < 0.002) (Figure 2D).

### Dominantly altered brain regions were indicative of subtype-specific rTMS targets

To identify subtype-specific rTMS targets, we assumed that the effective targets would be brain regions with prominent differentiation between depression subtypes and HC. We designed a novel method to identify rTMS targets based on brain-region level classifiers (Figure 3A). For each subtype, the top-10 brain regions whose ALFF values were most significantly discriminative from the HC were identified as candidate target regions; based on the dominant regions amongst, subtype-specific rTMS targets were identified.

**Figure 3.**
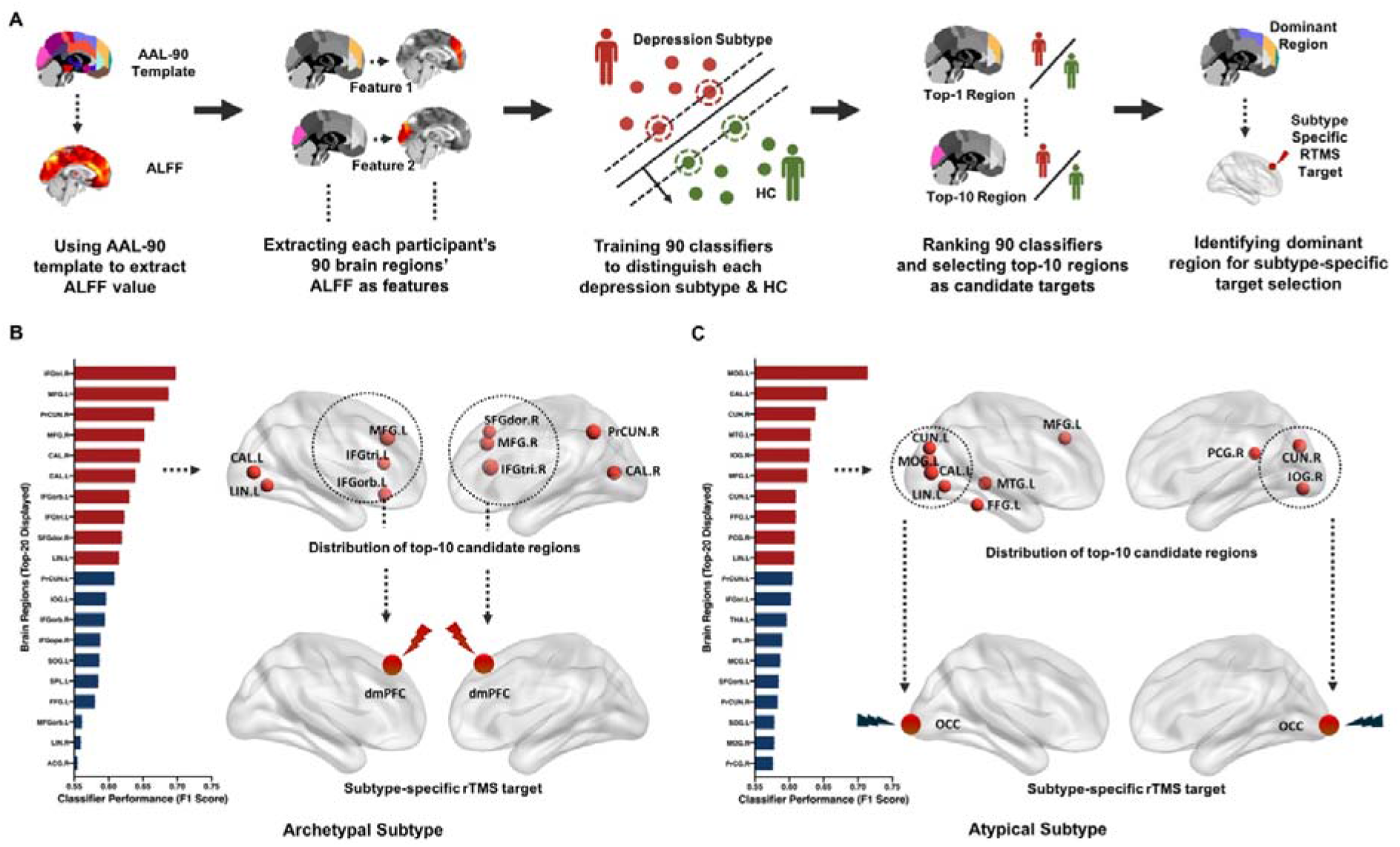
Dominantly altered brain regions were indicative of subtype-specific repetitive transcranial magnetic stimulation (rTMS) targets. Panel A shows the process of identifying subtype-specific rTMS targets. Panel B shows the top-20 brain-region F1 scores of the archetypal subtype; the top-10 candidate regions were dominantly distributed in the frontal region and dmPFC was suggested as subtype-specific rTMS target for the archetypal subtype (in medial view). Panel C shows the top-20 brain-region F1 scores of the atypical subtype; the top-10 candidate regions were dominantly distributed in the occipital region and OCC was suggested as subtype-specific rTMS target for the atypical subtype (in medial view). AAL=automated anatomical labeling; ALFF=amplitude of low frequency fluctuations; HC=healthy control; ACG.R=right anterior cingulate and paracingulate gyri; CAL.L= left calcarine fissure and surrounding cortex; CAL.R=right calcarine fissure and surrounding cortex; CUN.L=left cuneus; CUN.R=right cuneus; FFG.L=left fusiform gyrus; IFGope.R=right inferior frontal gyrus, opercular part; IFGorb.L= left inferior frontal gyrus, orbital part; IFGorb.R= right inferior frontal gyrus, orbital part; IFGtri.L=left inferior frontal gyrus, triangular part; IFGtri.R=right inferior frontal gyrus, triangular part; IOG.L=left inferior occipital gyrus; IOG.R=right inferior occipital gyrus; IPL.R=right inferior parietal, but supramarginal and angular gyri; LIN.L=left lingual gyrus; LIN.R=right lingual gyrus; MCG.L=left median cingulate and paracingulate gyri; MFG.L=left middle frontal gyrus; MFG.R=right middle frontal gyrus; MFGorb.L=left middle frontal gyrus, orbital part; MOG.L=left middle occipital gyrus; MOG.R=right middle occipital gyrus; MTG.L=left middle temporal gyrus; PCG.R=right posterior cingulate gyrus; PrCG.R=right precentral gyrus; PrCUN.L=left precuneus; PrCUN.R=right precuneus; SFGdor.R=right superior frontal gyrus, dorsolateral; SFGorb.L=left superior frontal gyrus, orbital part; SOG.L=left superior occipital gyrus; SPL.L=left superior parietal gyrus; THA.L=left thalamus; dmPFC=dorsomedial prefrontal cortex; OCC=occipital cortex.

For archetypal depression, candidate regions included the superior, middle, and inferior frontal gyri, precuneus, calcarine cortex, and lingual gyrus, which all exhibited significantly higher discriminative power. Since these regions were located predominantly in the bilateral frontal region (6 of the 10), dorsomedial prefrontal cortex (dmPFC) was selected as the rTMS target for archetypal depression instead of the conventional left dlPFC [23] (Figure 3B).

For atypical depression, candidate regions included the middle and inferior occipital gyri, calcarine cortex, cuneus, lingual gyrus, middle frontal gyrus, middle temporal gyrus, fusiform gyrus, and posterior cingulate gyrus. A bulk of these (6 of the 10) resided in the occipital region, so the occipital cortex (OCC) was selected as the rTMS target (Figure 3C).

### Precise rTMS normalized the frontal-posterior functional imbalance

Based on Center 1 findings, we deployed the neuroimaging-based subtyping and precise rTMS strategy to hospitalized, depressed youths at Center 2. One classifier was trained to first divide the depressed youths (n=72) into the archetypal subtype (n=16) and the atypical subtype (n=56). No significant differences in demographic or clinical characteristics were found between the two subtypes (Table 1). For archetypal depression, all 16 completed the treatment for the first week, and 10 completed the entire course. For atypical depression, all 56 completed the first-week treatment, and 41 completed the full course. No serious adverse events occurred in either subtype.

**Table 1.** Demographic and clinical information of hospitalized, depressed youths at Center 2.

| Variable | Archetypal Subtype<br>(N=16) |  | Atypical Subtype<br>(N=56) |  | T | P |
| --- | --- | --- | --- | --- | --- | --- |
|  | mean | SD | mean | SD |  |  |
| Age at hospitalization (years) | 16.38 | 2.25 | 16.05 | 2.39 | 0.48 | 0.63 |
| Age at onset (years) | 14.38 | 2.06 | 13.75 | 2.48 | 0.92 | 0.361 |
| Body mass index | 24.95 | 8.59 | 23.43 | 7.55 | 0.59 | 0.56 |
| **Baseline outcomes** |  |  |  |  |  |  |
| HAMD-17 | 25.25 | 5.85 | 24.73 | 4.34 | 0.39 | 0.70 |
| HAMA | 23.25 | 5.50 | 21.86 | 5.15 | 0.94 | 0.35 |
| | N | % | N | % | $\chi^2$ | P |
| Female | 10 | 62.50 | 42 | 75.00 | 0.45 | 0.50 |
| First episode | 4 | 25.00 | 29 | 51.79 | 3.60 | 0.06 |
| Suicidality | 12 | 75.00 | 44 | 78.57 | 0.00 | 1.00 |
| Non-suicidal self-injury | 9 | 56.25 | 43 | 76.79 | 1.69 | 0.19 |
| Previous medications | 13 | 81.25 | 40 | 71.43 | 0.22 | 0.64 |
| Antidepressant | 10 | 62.50 | 35 | 62.50 | 0.00 | 1.00 |
| Antipsychotic | 8 | 50.00 | 24 | 42.86 | 0.26 | 0.61 |
| Mood stabilizer | 3 | 18.75 | 17 | 30.36 | 0.36 | 0.55 |
| Sedative hypnotic | 3 | 18.75 | 9 | 16.07 | 0.00 | 1.00 |

SD=standard deviation; HAMD-17=17-items Hamilton Rating Scale for Depression;
HAMA=14-items Hamilton Rating Scale for Anxiety

For both subtypes, significant changes in frontal-posterior functional imbalance were observed after one week of treatment and were maintained throughout the treatment period (Figure 4A, Supplementary Table S1). In archetypal depression, significant ALFF decreases in frontal regions and significant ALFF increases in posterior regions were observed from T0 to T1 with sustained changes at T2, reversing the imbalance that was observed at the baseline. In atypical depression, converse changes in ALFF were detected with significantly increased ALFF in frontal regions and significantly decreased ALFF in posterior regions from T0 to T1. These changes were also sustained at T2. For both subtypes, ALFF changes from T1 to T2 were less significant, indicating the majority of changes occurred during the first week of rTMS (Supplementary Table S1).

**Figure 4.**
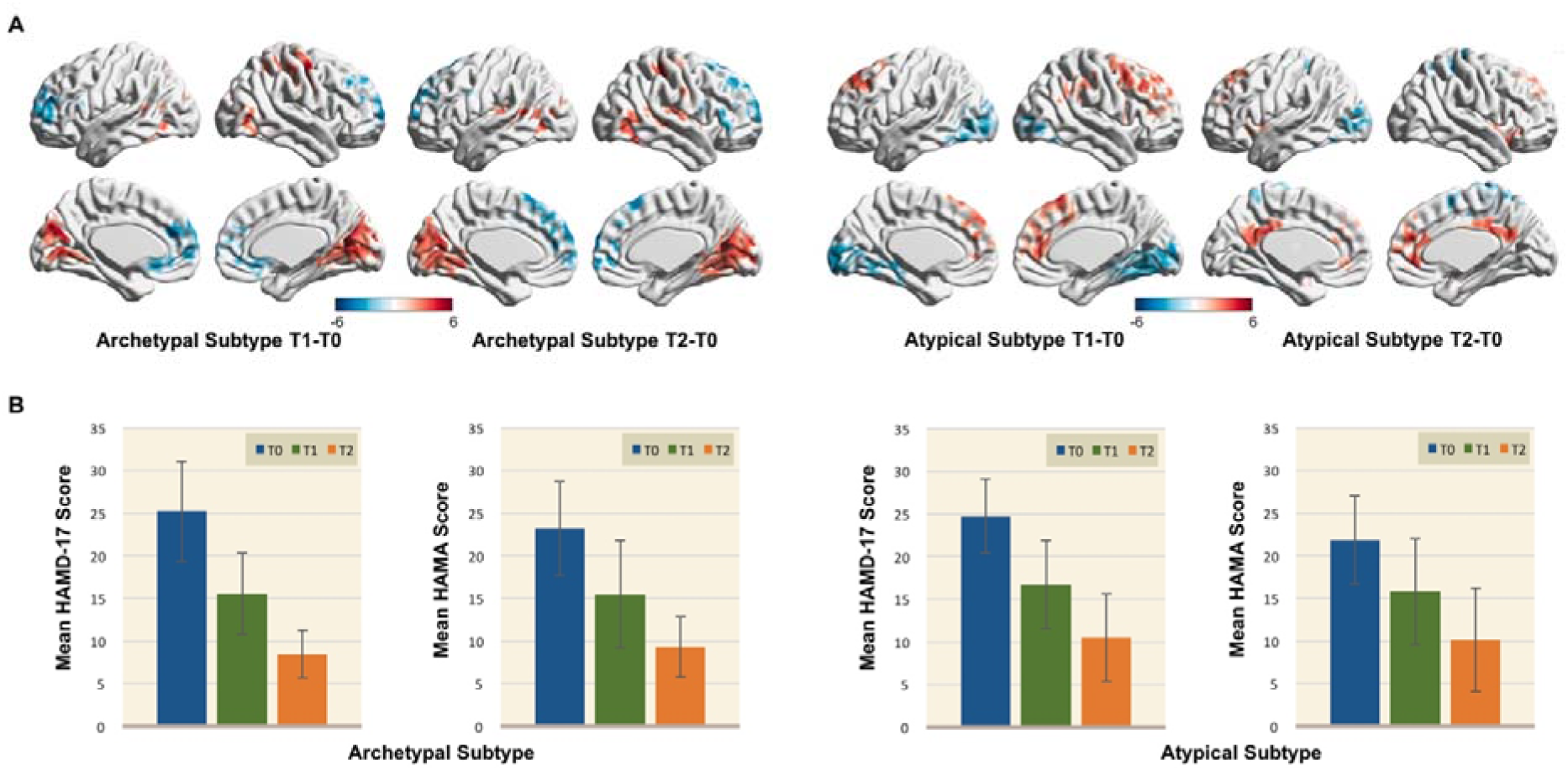
Neuroimaging and symptomatic improvements after precise repetitive transcranial magnetic stimulation (rTMS) in hospitalized, depressed youths. Panel A shows the neuroimaging alterations in both subtypes at midpoint T1 and terminal point T2 compared to baseline T0 after precise rTMS (whole-brain voxel-wise paired t-test of the amplitude of low-frequency fluctuations between T1 & T0 and T2 & T0 with voxel-level statistical significance p < 0.05, followed by Gaussian random field multiple-comparison correction; cluster-level p < 0.05, cluster size > 250). Post precise rTMS, distinct frontal-posterior functional imbalance in both subtypes were normalized. Red represented t > 0, blue represented t < 0. See Supplementary Table S1 for detailed brain regions. Panel B shows the mean HAMD-17 scores and mean HAMA scores of both subtypes at baseline T0, midpoint T1, and terminal point T2 after precise rTMS. Error bars represented standard deviation. HAMD-17=17-item Hamilton Depression Rating Scale; HAMA= Hamilton Anxiety Rating Scale.

### Precise rTMS showed great clinical efficacy

We observed significant symptomatic improvements in both subtypes of depressed youths after precise rTMS. The primary outcome was symptom severity measured by both the HAMD-17 and HAMA since depressive and anxiety symptoms are highly co-occurring in depressed youths [32, 33]. Changes in HAMD-17 and HAMA scores corresponded with ALFF changes from baseline T0 to T2 (Figure 4B). For the primary outcome, 31.25% of the archetypal subtype and 28.57% of the atypical subtype had an overall response at T1; 90.00% of the archetypal subtype and 70.73% of the atypical subtype exhibited an overall response at T2. Remission was achieved in 6.25% of the archetypal subtype and 5.36% of the atypical subtype at T1, which increased to 30.00% and 46.34%, respectively, at T2 (Supplementary Table S2).

For the secondary outcome of suicidality, 12 patients in the archetypal subtype and 44 in the atypical subtype had a baseline score > 0 on HAMD-17 Item-3. At T1, 83.33% of the archetypal subtype and 65.91% of the atypical subtype had a response, which increased to 100.00% and 81.25% at T2. Remission was achieved in 58.33% of the archetypal subtype and 38.64% of the atypical subtype at T1; and 100.00% and 56.25%, respectively, at T2 (Supplementary Table S2).

## DISCUSSION

In this study, we identified an objective neurobiomarker to guide subtyping and precise treatment in depression, bridging critical translational gaps in actualizing precision medicine in Psychiatry. We developed a neuroimaging-based subtyping and precise rTMS strategy for depression at Center 1 and implemented it in hospitalized, depressed youths with pre- and post-treatment neuroimaging and symptom assessments at Center 2. Using machine learning techniques, we identified two depression subtypes based on ALFF patterns at Center 1, namely archetypal and atypical depression. The subtypes found herein replicated our prior findings [15], further implicating frontal-posterior functional imbalance as a reproducible neurobiomarker for depression. Moreover, we identified dmPFC and OCC as subtype-specific rTMS targets for the archetypal and atypical depressions, respectively. We next subtyped the hospitalized, depressed youths at Center 2 through a classifier we trained in Center 1 and delivered precise rTMS. Excitingly, the observed patterns of ALFF alterations in both archetypal and atypical depression subtypes appeared to normalize in response to rTMS of subtype-specific targets with corresponding improvement in depressive and anxiety symptoms, and suicidality.

Depression is highly heterogeneous with varying etiologies due to its diagnostic criteria relying primarily on descriptive symptoms [8]. Several attempts have been made to delineate its heterogeneity based on neuroimaging features, which have the potential of unveiling neural mechanisms of psychiatric disorders. However, the reliability of identified neuroimaging subtypes has been limited by the lack of cluster reproducibility [24], proper validation [34], or clinical ramification [35]. Despite that no answer exists for universally applicable clustering due to the nature of clustering and the lack of class labels, to meet the urgent need in depression, we suggest an appropriate subtyping for depression would be identified by objective neurobiomarkers capable of guiding precision medicine [36].

Intriguingly, we identified two depression neuroimaging subtypes with high stability and reproducibility: archetypal depression, which had significantly increased ALFF in frontal regions and significantly decreased ALFF in posterior regions compared to HC, and atypical depression, which showed a converse pattern of ALFF alterations. Consistent with our previous finding in a nonoverlapping sample [15], the archetypal and atypical depression subtypes showed converse patterns of frontal-posterior functional imbalance, suggesting the functional hierarchy between higher-order, heteromodal areas and primary, unimodal cortices as a latent mechanism of depression [37]. Moreover, neurostimulation of frontal and posterior cortices could be transmitted through their functional connectome and accordingly modulate regional neuroimaging deficits [38]. Normalization of the frontal-posterior functional imbalance following rTMS in the depressed youths herein supports strong promise for our neuroimaging-based subtyping in clinical practice. Further studies are warranted to confirm the translational implications of the depression subtypes identified by objective neurobiomarkers.

The dmPFC was selected as the rTMS target for archetypal depression. It has significant connections to sensorimotor areas and the anterior cingulate cortex, serving as a conduit between cognitive control and emotional processing regions [39]. Historically, the dlPFC was selected as the conventional rTMS target due to its potential pathophysiological modulation of depression [40]. However, convergent studies in lesion, stimulation and neuroimaging suggested that the dmPFC was not only the most promising target alternative to the dlPFC but also played a more central role in the underlying neurophysiology of depression, as was proposed by Downar et al. [41]. Our findings further supported the dmPFC as an effective rTMS target, which was identified based on objective neuroimaging measures rather than based on purely empirical findings. Conversely, the OCC was identified as the rTMS target for atypical depression. The OCC is a primary sensory region responsible for visual processing [42]. Decreased gamma-aminobutyric acid (GABA) in the OCC has been previously observed in depressed patients [43]. Further, it was found to be associated with the altered visual perception that correlated with symptom severity in acute MDD [44]. Decreased GABA may underlie neural mechanisms for increased ALFF in posterior regions in atypical depression [45]. Of note, both human and animal neuroimaging studies have observed excitatory effects of high-frequency rTMS and inhibitory effects of low-frequency rTMS on OCC [38, 46, 47]. Hence, in attempt to dampen the increases in posterior ALFF, we delivered low-frequency rTMS to the OCC in atypical depression. Altogether, the dmPFC and OCC appear to be subtype-specific rTMS targets with strong translational implications, bolstered by prior neurophysiological findings. Our novel strategy for precise rTMS in hospitalized, depressed youths achieved reasonably high efficacy based on both neuroimaging and symptom outcomes. The normalization of ALFF patterns in both subtypes strongly supported the importance of frontal-posterior functional imbalance as an objective biomarker to guide precision medicine in depression. Given the high concurrence of depression and anxiety during adolescence and early adulthood [32, 33], we used depressive and anxiety symptoms in determining the primary outcome. Due to limited studies in depressed youths, there is no consensus on the efficacy of conventional rTMS targeting the dlPFC. However, conventional rTMS generally have response rates of 29-46% and remission rates of 18-31% in depression [23]. In this study, we achieved higher response rates of 90.00% and 70.73% in archetypal and atypical depression, respectively, as well as respective remission rates of 30.00% and 46.34%. Furthermore, consistent with conventional rTMS studies [48], our precise rTMS also reduced suicidality in depressed youths. While not strictly meeting the criteria of TRD, our hospitalized, depressed youths had complicated clinical features (see Supplementary Material) that could define the group as having difficult-to-treat depression [49]. In short, our precise rTMS strategy significantly outperformed conventional rTMS in depressed youths.

The current study may be improved in several ways. Sham-controls were not applied due to the exploratory nature of our rTMS trial and our belief that all hospitalized patients deserved active treatment. Further, participants were not blinded to their rTMS targets, and placebo effects may confound the findings. In addition, the sample size of our rTMS trial was relatively small. Larger, double-blinded, and randomized controlled trials are needed to confirm the clinical implications of our neuroimaging-based framework. Longer treatment courses may also be warranted to improve clinical efficacy and increase remission rates. The high response but comparatively low remission rates may indicate the need for extended treatment beyond two weeks.

In summary, we developed a novel precision medicine framework for depression that went beyond symptomatic measures and incorporated neuroimaging-based subtyping to guide precise rTMS with promising results. Further studies are warranted in larger depressed samples and for longer durations, as well as using a double-blinded, randomized controlled design.

## Data Availability

All data produced in the present work are contained in the manuscript

## ACKNOWLEDGEMENTS

The authors were supported by National Science Fund for Distinguished Young Scholars (81725005 to Fei Wang), National Natural Science Foundation Regional Innovation and Development Joint Fund (U20A6005 to Fei Wang), Jiangsu Provincial Key Research and Development Program (BE2021617 to Fei Wang), National Natural Science Foundation of China (62176129 to Xizhe Zhang), Key Project supported by Medical Science and Technology Development Foundation, Jiangsu Commission of Health (ZD2021026 to Rongxin Zhu), Longer Life Foundation (2017-005 to Fay Y. Womer), and Hong Kong Global STEM Professor Scheme (to Weixiong Zhang).

