## Supplementary Material for "A Neuroimaging-based Precision Medicine Framework for Depression"

### METHODS

#### *Participants*

Diagnoses: patients 18 years of age and older were diagnosed through the Structured Clinical Interview for DSM-IV, SCID; patients younger than 18 years old were diagnosed through the Schedule for Affective Disorders and Schizophrenia for School-Age Children-present and Lifetime Version, K-SADS-PL.

Detailed exclusion criteria: major medical condition (e.g., high blood pressure, diabetes, cancer, etc.); neurological disorder (e.g., epilepsy, brain cyst, space occupying brain lesion, significant head trauma with loss of consciousness for longer than five minutes etc.); MRI contraindications (e.g., claustrophobia, non-removable metal implants, etc.); excessive head motion during MRI scanning (more than 3.0° rotation and 3.0 mm translation).

For participants in Center 1, 238 major depressive disorder (MDD) outpatients (mean age: 24.13 ± 6.81; sex: 68 male/170 female) and 66 healthy controls (HC) (mean age: 25.18 ± 7.14; sex: 14 male/52 female) recruited from April 2019 to May 2020 were included in the study. No significant differences of mean age or sex were found between patients and HCs (age: t=-1.10, p=0.27; sex: $\chi^2$=1.42, p=0.23). Patients in Center 2 were 72 youths (mean age: 16.13 ± 2.35; sex: 20 male/52 female) hospitalized from September 2020 to April 2021. Sex was ascertained by self-report. All the participants were Chinese.

The hospitalized, depressed youths recruited from Center 2 presented with complicated clinical features. Among the 72 patients who were in a major depressive episode, 38 patients were diagnosed as MDD and 34 patients were diagnosed as bipolar II disorder (BD II). The age at their initial onset of depression ranged from 9 to 20, with an average of 13.89 ± 2.39 years old; the overall duration varied from half a month to 108 months, and the mean length was 26.02 ± 20.09 months. Thirty three patients were in their first major depressive episode, while 39 patients experienced at least two episodes (2 episodes: 10 patients; 3 episodes: 7 patients; 4 episodes: 1 patient; 5 episodes or more: 21 patients). For the current episode, 30 patients had a heterogeneous clinical presentation and should be described with certain specifiers; 22 patients therein were with mixed features, seven patients were with psychotic features and one patient was with atypical features.

For those who had recurrent major depressive episodes, 17 patients could be described with the longitudinal course specifier, namely without full inter-episode recovery, while four patients met the criteria of rapid-cycling specifier. Besides, we also captured a subclinical inner-episode feature among the hospitalized young patients that we characterized as “waxing and waning”; specifically, during one major depressive episode, the patient would experience frequent onsets of severe depression, each followed by a short period of recovery which sometimes could only last for a day or two. Thirty four patients revealed this feature, and 29 of them were in their first episode. Altogether, we suggested that the hospitalized, depressed youths, with either single-episode or recurrent episodes, were lack of clinically significant recovery, which consequently led to an inner-

or inter-episode waxing and waning of the course. As a result, all the patients reported serious impairment in academic and social functioning, coupled with a high risk of self-harm.

***MRI data acquisition and processing***

Center 1: Participants received a resting-state functional MRI (fMRI) scanning on an GE Discovery MR750W 3.0 T scanner after assessment for eligibility. The scanning lasted eight minutes using gradient echo planar imaging (GRE-EPI) sequence; the parameters are listed below: TR=2000ms, TE=30ms, FA=90 °, FOV=220*220mm$^2$, matrix=64*64, slices=36, thickness=4mm, gap=0mm, volume=240, voxel size=3.4*3.4*4mm$^3$.

Center 2: For each patient, MRI scanning were performed at three time points: baseline T0 (one day before precise repetitive transcranial magnetic stimulation, rTMS), midpoint T1 (after one-week precise rTMS) and terminal point T2 (after two-week precise rTMS). The imaging data were obtained from a Siemens MAGNETOM Prisma 3.0T scanner. The resting-state fMRI scanning lasted for eight minutes and seven seconds using GRE-EPI sequence while the structural MRI scanning lasted for five minutes and 58 seconds using magnetization-prepared rapid acquisition gradient echo (MPRAGE) sequence; the parameters are listed below. Resting-state fMRI: TR=500ms, TE=30ms, FA=60°, FOV=224*224mm$^2$, matrix=64*64, slices=35, thickness=3.5mm, gap=0.5mm, volume=960, voxel size=3.5*3.5*3.5mm$^3$. T1-weighted structural images: TR=2530ms, TE=2.98ms, FA=7°, FOV=224*256mm$^2$, matrix=256*256, slices=192, thickness=1mm, voxel size=0.5*0.5*1.0mm$^3$.

The resting-state fMRI data were processed using the Statistical Parametric Mapping 8 (SPM8; http://www.fil.ion.ucl.ac.uk/spm) and Data Processing Assistant for Resting-State fMRI (DPARSF; http://www.restfmri.net/forum/DPARSF) toolkits. The preprocessing procedures are listed as below: removing the first 10 time points; slice-timing correction; realignment and head-motion correction; normalization to standard EPI template in Montreal Neurological Institute (MNI) space and resample to 3*3*3 mm$^3$; spatial smoothing with a 6-mm full width at half maximum Gaussian kernel; linear detrending; temporal band-pass filtering (0.01-0.08 Hz); for more details on preprocessing, see our previous study [1]. After preprocessing, amplitude of low frequency fluctuations (ALFF) [2] was calculated as the index. For each voxel, the square root of the power spectrum in the low-frequency range was suggested as its ALFF value; then it was divided by the global mean ALFF value for standardization. The automated anatomical labeling (AAL) 90 atlas [3] was applied to filter the ALFF data, leaving 47636 voxels for each subject.

***Identifying neuroimaging subtypes of depression***

Firstly, the high-dimensional AAL-filtered ALFF data of MDD patients were reduced to 2-dimensional while preserving the structure of the original neighboring points through the t-distributed Stochastic Neighbor Embedding (t-SNE) algorithm [4]; the perplexity was set to 30 and the learning rate was set to 200. Secondly, the low-dimensional data were divided into two subtypes through the Gaussian Mixture Model (GMM) algorithm, whose performance has been shown to be superior in previous studies [5]. The type of covariance parameters was set to “full” to ensure each component had its own general covariance matrix and the convergence threshold was set to 1e-3. The stability of depression subtypes was tested based on the Consensus Clustering method [6]; 80% subjects were randomly selected and subtyped each time, and the process of which was repeated for 500 times. The consensus matrix $M$ was a $(N * N)$ matrix defined as followed ($N$ represented the number of patients):

$$M(i,j) = \frac{\sum_{h=1}^{500} M^{(h)}(i,j)}{\sum_{h=1}^{500} I^{(h)}(i,j)}$$

Specifically, $M^{(h)}$ denoted the connectivity matrix corresponding to the iteration $h$, such that its $(i,j)$- *th* entry was equal to 1 if patients $i$ and $j$ belonged to the same subtype, and 0 otherwise. $I^{(h)}$ denoted the indicator such that its $(i,j)$-*th* entry was equal to 1 if both patients $i$ and $j$ were present in the iteration $h$, and 0 otherwise. Further, the subtype stability $s_i$ could be denoted as $m(s_i)$ , representing the average score of all pair of patients who belonged to the same subtype in the consensus matrix $M$ :

$$m(s_i) = \frac{1}{N_{s_i}(N_{s_i} - 1)/2} \sum_{\substack{i,j \in I_{s_i} \\ i<j}} M(i,j)$$

Next, using the filtered ALFF data as features, Support Vector Machine (SVM) classifiers were trained for distinguishing MDD subtypes and total MDD sample from HC. Specifically, the Radial Basis Function (RBF) was employed as the kernel function for the SVM classifiers, with the kernel coefficient setting to reciprocal of features number and the regularization parameter setting to 10. The area under the curve (AUC) and F1 score were used for evaluating classifier performance, and further verified through a five-fold cross-validation procedure. The receiver operating characteristic (ROC) curves were plotted for comparison. Permutation testing was used to further evaluate reliability of depression subtypes [7]. For patients in each subtype, the labels were randomly permuted into either archetypal or atypical; SVM classifier was trained using the same protocol as mentioned above and the F1 score was calculated, following five-fold cross-validation. For each subtype, the procedure was repeated for 500 times. Statistical significance level was set to $p < 0.002$; when true F1 scores exceeded 95% confidence interval of permutated F1 scores, the result of permutation testing was deemed significant. The scikit-learn 0.22.2.post1 python library was used in our data analysis [8].

***Identifying subtype-specific RTMS targets***

First, we extracted the voxel-wise ALFF values of the 90 brain regions based on AAL 90 atlas and each brain region was extracted as neuroimaging features. Next, SVM classifiers were trained using the features extracted from each brain region to distinguish each depression subtype from HC; as such, a total of 90 classifiers, one for each brain region, were trained for every depression subtype. The training protocol of the classifiers was the same as that mentioned above. The F1 score was taken as the indicator to assess the performance of each classifier, followed by a five-fold cross-validation procedure; algorithms applied were from the scikit-learn 0.22.2.post1 python library [8].

Consequently, we ranked the 90 brain regions based on the F1 score and selected the top 10 regions in each subtype as candidate regions. For each depression subtype, the anatomical distributions of 10 candidate regions were integrated; major brain region component was regarded as the dominant region, combined with current neurobiological understanding we would determine subtype-specific rTMS target.

***Transcranial magnetic stimulation protocol***

A Magneuro 60 magnetic stimulator (VISHEE Inc., Nanjing, China) was used to deliver the rTMS treatment. Patients received rTMS treatment twice a day and 5 consecutive days per week; on treatment days, at least a one-hour interval between two sessions was required. For the archetypal subtype, the stimulation was high frequency (10Hz), with 40 pulses per train; 30 trains with a 10-second interval were delivered; in total, there were 1,200 pulses per session. For the atypical subtype, the stimulation was low frequency (1Hz), with 10 pulses per train; 100 trains with a 2-second interval were delivered; in total, there were 1,000 pulses per session.

***Precise localization***

Depressed youths received precise rTMS target localization. The steps are listed below. We integrated the international 10-10 system and individualized three-dimensional MRI for joint positioning so the target region could be localized more accurately. Since the scalp location for targeting dorsomedial prefrontal cortex (dmPFC) corresponds to 25% of the nasion-inion distance [9], we chose the AFz label, which corresponds to 20% of the nasion-inion distance, as the coordinate reference in the 10-10 system for targeting dmPFC in the archetypal subtype. Meanwhile, the Oz label located on the scalp of the occipital lobe was suggested as the coordinate reference for the atypical subtype [10].

Before MRI scanning, every patient was equipped with a formfitting vinyl head cap, the surface of which was fixed with two cod liver oil capsules on both AFz and Oz reference points. The patient then underwent MRI scanning, during which the individualized anatomical structures of both dmPFC and occipital cortex (OCC) were localized through the sagittal plane using T1-weighted images. To localized dmPFC, two tangent lines along both the trunk and the genu of corpus callosum were firstly drawn; their intersection, located generally around the border of the prefrontal cortex and the dorsal anterior cingulate cortex, was used as the initial point on which to project a ray at the closest distance to the cortex. The projection point on the scalp was finally indicated as the patient's individualized anatomical dmPFC localization. On the other hand, the midpoint of the outer edge along the cuneus was suggested as the individualized anatomical OCC localization. The distances between AFz-dmPFC and between Oz-OCC were then measured and recorded accordingly. After MRI scanning, patients were divided into either archetypal or atypical neuroimaging subtype; by means of the subtype label, we could firstly choose the appropriate 10-10 system reference point (AFz/OZ). Next, the patient wore the same vinyl head cap that he/she wore when going through MRI scanning; the coil should be initially placed on the 10-10 system reference point, then removed by the same distance as was recorded between its anatomical target and reference point. Using the procedures mentioned above, the individualized target regions of both subtypes could be localized precisely.

**Supplementary Table S1.** ***Paired t-test post-pre precise repetitive transcranial magnetic stimulation (rTMS).***

| Brain Region | Peak MNI Coordinate | | | Peak Intensity | Number of Voxels |
|---|---|---|---|---|---|
| **T1-T0** | | | | | |
| **Archetypal subtype** | | | | | |
| Left middle frontal gyrus | -39 | 48 | 24 | -6.34 | 1663 |
| Left calcarine fissure and surrounding cortex | -3 | -66 | 18 | 6.22 | 2616 |
| Right precentral gyrus | 45 | -9 | 57 | 5.56 | 636 |
| **Atypical subtype** | | | | | |
| Left middle occipital gyrus | -21 | -93 | 3 | -5.05 | 2763 |
| Right superior frontal gyrus, medial | 6 | 27 | 63 | 5.40 | 2849 |
| Right inferior parietal, but supramarginal and angular gyri | 57 | -30 | 51 | 4.07 | 290 |
| **T2-T0** | | | | | |
| **Archetypal subtype** | | | | | |
| Left fusiform gyrus | -39 | -54 | -12 | 7.11 | 3162 |
| Left superior frontal gyrus, dorsolateral | 33 | 51 | 36 | -9.94 | 1786 |
| Right postcentral gyrus | 39 | -39 | 60 | 4.95 | 347 |
| **Atypical subtype** | | | | | |
| Left middle occipital gyrus | -42 | -87 | 0 | -4.84 | 400 |
| Right caudate nucleus | 18 | 3 | 18 | 4.54 | 484 |
| Left caudate nucleus | -15 | 9 | 15 | 5.21 | 480 |
| Right anterior cingulate and paracingulate gyri | 3 | 45 | 21 | 5.20 | 913 |
| Left posterior cingulate gyrus | 0 | -36 | 30 | 4.97 | 488 |
| Right supplementary motor area | 9 | 3 | 63 | -4.40 | 764 |
| **T2-T1** | | | | | |
| **Archetypal subtype** | | | | | |
| None | | | | | |
| **Atypical subtype** | | | | | |
| Right precentral gyrus | 30 | -6 | 48 | 4.90 | 1116 |

**Supplementary Table S2.** ***Response rate and remission rate of the primary and secondary outcome.***

| Outcome | Archetypal Subtype | | Atypical Subtype | |
|---|---|---|---|---|
| **Primary Outcome** | **N** | **%** | **N** | **%** |
| **Week 1** | **N=16** | | **N=56** | |
| **HAMD-17** | | | | |
| Response | 4 | 25.00 | 14 | 25.00 |
| Remission | 1 | 6.25 | 1 | 1.79 |
| **HAMA** | | | | |
| Response | 3 | 18.75 | 10 | 17.86 |
| Remission | 1 | 6.25 | 3 | 5.36 |
| **Overall** | | | | |
| Response | 5 | 31.25 | 16 | 28.57 |
| Remission | 1 | 6.25 | 3 | 5.36 |
| **Week 2** | **N=10** | | **N=41** | |
| **HAMD-17** | | | | |
| Response | 8 | 80.00 | 29 | 70.73 |
| Remission | 3 | 30.00 | 11 | 26.83 |
| **HAMA** | | | | |
| Response | 8 | 80.00 | 23 | 56.10 |
| Remission | 2 | 20.00 | 18 | 43.90 |
| **Overall** | | | | |
| Response | 9 | 90.00 | 29 | 70.73 |
| Remission | 3 | 30.00 | 19 | 46.34 |
| **Secondary Outcome** | **N** | **%** | **N** | **%** |
| **Week 1** | **N=12** | | **N=44** | |
| **Suicidality** | | | | |
| Response | 10 | 83.33 | 29 | 65.91 |
| Remission | 7 | 58.33 | 17 | 38.64 |
| **Week 2** | **N=6** | | **N=32** | |
| **Suicidality** | | | | |
| Response | 6 | 100.00 | 26 | 81.25 |
| Remission | 6 | 100.00 | 18 | 56.25 |

HAMD-17=17-items Hamilton Rating Scale for Depression; HAMA=14-items Hamilton Rating Scale for Anxiety; Overall=HAMD-17 or HAMA; suicidality=3rd item of HAMD-17; response: $\geq$ 50% reduction from the baseline score; HAMD-17 remission: score $\leq$ 7; HAMA remission: score $\leq$ 7; suicidality remission: score=0
